# An update to the critical appraisal of Milwaukee protocol

**DOI:** 10.1101/2022.12.14.22283490

**Authors:** Ferhat Arslan, Haluk Vahaboglu

## Abstract

Rabies is a highly fatal encephalitis. Currently, there is no approved treatment. Inducing therapeutic coma during the first week of symptomatic rabies patient, called Milwaukee protocol, had been suggested as promising. However, recent evidence failed to support the use of the Milwaukee protocol. This mini-review analyzed the reports of patients managed with therapeutic coma since 2014 to provide an update for the critical appraisal of this protocol.

## Introduction

Rabies is an almost uniformly fatal encephalitis caused by the members of the genus Lyssavirus. During the last thirty years, the global burden of rabies is declined due to increased living conditions and the availability of vaccines in rural areas (1). Bats and infected dogs are the primary vehicles of human rabies. Two clinical forms are described as paralytic and furious, the latter even more dramatic (2). The virus enters the axons from neuromuscular junctions after entering through broken skin or mucosa. Viruses are transferred to the dorsal root ganglia and, with secondary transport, reach the brain (3). Naturally, in a maximum of 14 days from the first symptoms appear, patients die due to respiratory or cardiac arrest (4).

The pathophysiology of neuronal damage is yet to be elucidated. Autopsy findings of patients who died of rabies vary; some show extensive damage to the brain tissue (5), while others do not (6). However, animal studies displayed extensive neuronal damage attributed to mitochondrial dysfunction and apoptosis (7, 8). Despite the progress in understanding the pathophysiology of neuronal damage in rabies, approved treatment has not yet been developed (9). In 2005 a 15-year-old girl developed rabies-like encephalitis after bat exposure and survived with the experimental Milwaukee protocol (10). The Milwaukee protocol recommends inducing therapeutic coma by ketamine and midazolam during rabies patients’ first week of ICU admittance (8). Despite the great excitement of this first report, Milwaukee protocol failed to replicate the initial success during the following years. It has been proposed to abandon therapeutic coma induction during ICU care of rabies patients (8, 11). However, since the first warning in 2015 to leave inducing therapeutic coma in rabies management has been ignored, the Milwaukee protocol is still being used (8).

This mini-review aimed to recruit survival evidence on Milwaukee protocol since 2015.

## Data collection

We screened the Web of Science using “rabies (All fields) AND Milwaukee (All fields),” refined dates from 2015 to 2022, carefully inspected abstracts and identified data for 13 cases (Table 1). Next, we screened the Web of Science using “rabies (All fields) AND survived (All fields) AND case OR patient (All fields) to identify survived cases since 2015 who have not managed with induced therapeutic coma.

**Table 1.** Characteristics of rabies cases applied Milwaukee protocol since 2015

|  | Age/gender | Year<sup>1</sup> | Died of | Hosp. stay (days) | PEP | outcome | LAMA | ref |
| --- | --- | --- | --- | --- | --- | --- | --- | --- |
| 1 | 33/M | 2021 | Uncontrolled hypotension | 18 | none | Died |  | (14) |
| 2 | 27/F | 2021 | Respiratory failure | 580 | 5x RABV | Died |  | (15) |
| 3 | 51/M | 2021 | Not available | 7 | Ig & 5x RABV | Died |  | (16) |
| 4 | 29/M | 2020 | Not stated | 28 | 5x RABV | Died |  | (17) |
| 5 | 58/F | 2020 | Not stated | 17 | 3x RABV | Died | yes | (17) |
| 6 | 34/M | 2020 | Not stated | 2 | 3x RABV | Died |  | (17) |
| 7 | 12/M | 2019 | Autonomic dysregulation | 15 | none | Died |  | (18) |
| 8 | 58/F | 2017 | Heart block, profound acidosis | 8 | none | Died |  | (6) |
| 9 | 10/M | 2016 | Not stated | 4 | none | Died |  | (19) |
| 10 | 9/F | 2015 | Refractory hypotension | 75 | none | Died |  | (5) |
| 11<sup>2</sup> | 8/F | 2015 |  |  | none | Survived |  | (20) |

<sup>1</sup> Publication year
<sup>2</sup> Cat scratch

## Results

Table 1 outlines the characteristics of 12 cases managed with induced therapeutic coma published after 2014 in the medical literature. Briefly, only one patient survived (patient 12). This patient, however, was applied to the hospital with atypical symptoms such as acute abdominal pain, sore throat, and a mild cough. The initial diagnosis of patient 12 was aspiration pneumonia, and the exposure was a cat scratch. The diagnosis was made with anti-rabies antibody titration, and rabies antigen or RNA has never been shown. Patient 12 was managed under induced coma for ten days, and following the cessation of therapeutic coma patient gradually improved and was finally discharged with minimal neurologic deficit. Case reports stated the death reasons for patients 1, 2, 7, 8, and 10 (Table 1). Death among patients with therapeutic coma mostly was due to respiratory and cardiac failure. Interestingly, nosocomial infections were not reported if they ever occurred. Six patients received post-exposure prophylaxis (PEP) 3-5 times, and one patient received both rabies immunoglobulin (Ig) and a full course of rabies vaccine (RABV).

Table 2 displays patients managed with conventional ICU care and survived. Three dead patients were also included in the table because these were presented in the case series with surviving patients (12). A total of eight patients survived; all had 3-5 PEP, and two had post-exposure Ig. The follow-up period of survived patients was a minimum of six months and a maximum of four and a half years.

**Table 2.**
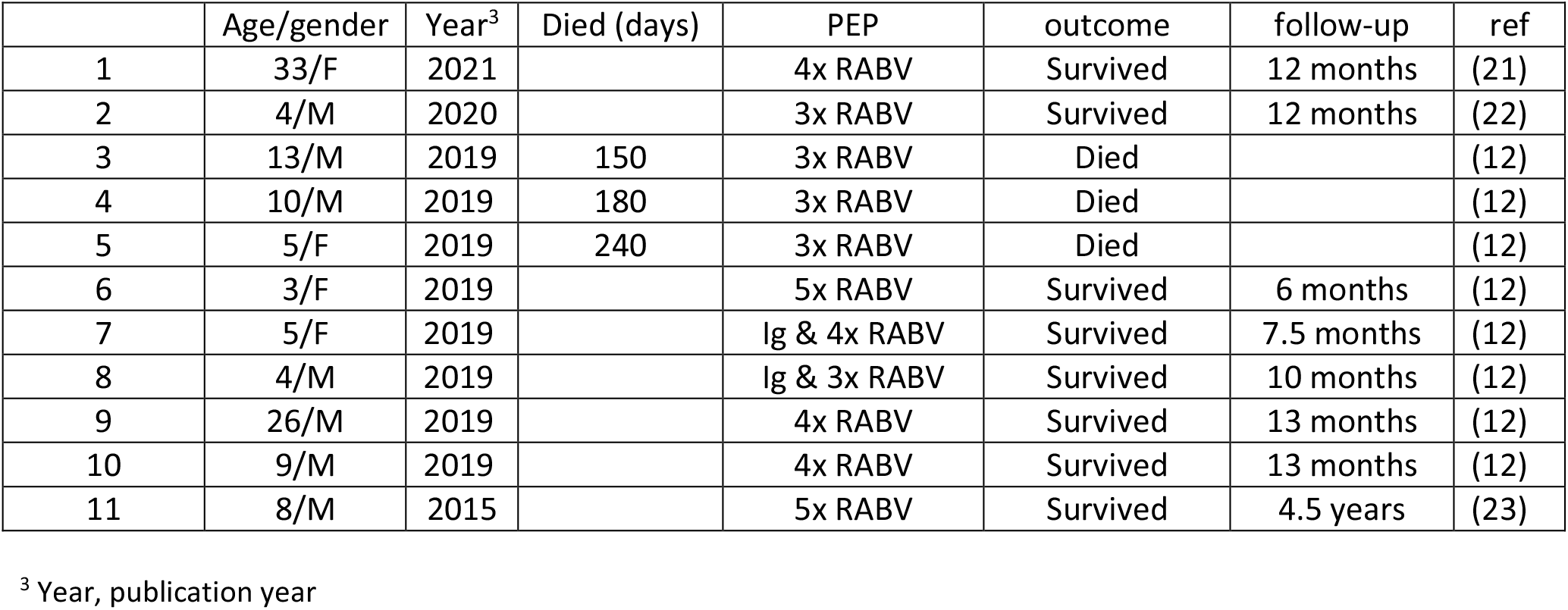
Characteristics of rabies cases not applied Milwaukee protocol since 2015 and survived

## Discussion

This mini-review aimed to address whether all rabies survivors had managed through induced therapeutic coma and found that this is not the case. Although we found one patient survived with Milwaukee protocol management, the diagnosis for that case was not robust. The patient was admitted with pneumonia. Moreover, she improved gradually after ending the therapeutic coma. The authors did not discuss what would happen if they had completed the therapeutic coma before ten days or even not inducing it at all.

The induced therapeutic coma may cause severe side effects such as electrolyte disturbances and cardiorespiratory arrest (13). Potential side effects, including facilitating the acquisition of nosocomial infections and causing immune depression, were discussed previously (8). Our mini-review also provided evidence that some patients died of nosocomial infections acquired under therapeutic coma. Uncontrolled hypotension, acidosis, and respiratory failure might be because of sepsis and, eventually, septic shock developed during the therapeutic coma. However, the authors neither gave any information nor discussed nosocomial infections.

The definitive diagnosis of rabies is almost always difficult in the early course of the disease (4). Therefore, applying potentially hazardous and experimental Milwaukee protocol, namely the therapeutic coma, might also harmful to patients with other mimicking encephalitides.

Finally, we agree with the authors who previously warned about abandoning Milwaukee protocol (inducing therapeutic coma) in the management of rabies (8, 11).

## Data Availability

All data produced in the present work are contained in the manuscript

## Notes

### Competing Interest Statement

The authors have declared no competing interest.

### Funding Statement

This study did not receive any funding

